# Large Language Models in Radiology Reporting—A Systematic Review of Performance, Limitations, and Clinical Implications

**DOI:** 10.1101/2025.03.18.25324193

**Authors:** Yaara Artsi, Eyal Klang, Jeremy D. Collins, Benjamin S. Glicksberg, Panagiotis Korfiatis, Girish N Nadkarni, Vera Sorin

**Affiliations:** Azrieli Faculty of Medicine, Bar-Ilan University, Zefat, Israel; The Charles Bronfman Institute of Personalized Medicine, Icahn School of Medicine at Mount Sinai, New York, New York, USA; The Windreich Department of Artificial Intelligence and Human Health, Mount Sinai Medical Center, NY, USA; The Hasso Plattner Institute for Digital Health at Mount Sinai, Icahn School of Medicine at Mount Sinai, New York, NY, USA; Department of Radiology, Mayo Clinic, Rochester, MN, USA

**Keywords:** Radiology reports, Large Language Models, Artificial Intelligence, Natural Language Processing, Automated Reporting, Clinical Evaluation, AI Alignment, Generative AI

## Abstract

**Background:** Large language models (LLMs) have emerged as potential tools for automated radiology reporting. However, concerns regarding their fidelity, reliability, and clinical applicability remain. This systematic review examines the current literature on LLM-generated radiology reports.

**Methods:** We conducted a systematic search of MEDLINE, Google Scholar, Scopus, and Web of Science to identify studies published between January 2015 and February 2025. Studies evaluating LLM-generated radiology reports were included. The study follows PRISMA guidelines. Risk of bias was assessed using the Quality Assessment of Diagnostic Accuracy Studies (QUADAS-2) tool.

**Results:** Nine studies met the inclusion criteria. Of these, six evaluated full radiology reports, while three focused on impression generation. Six studies assessed base LLMs, and three evaluated fine-tuned models. Fine-tuned models demonstrated better alignment with expert evaluations and achieved higher performance on natural language processing metrics compared to base models. All LLMs showed hallucinations, misdiagnoses, and inconsistencies.

**Conclusion:** LLMs show promise in radiology reporting. However, limitations in diagnostic accuracy and hallucinations necessitate human oversight. Future research should focus on improving evaluation frameworks, incorporating diverse datasets, and prospectively validating AI-generated reports in clinical workflows.

## Introduction

Radiology reports are the main communication interface between radiologists and referring physicians [1]. They can significantly influence medical decisions. However, there are numerous challenges associated with current radiology reporting that require attention.

One challenge in radiology reporting is inter-observer variability, referring to differences in how radiologists describe and interpret the same imaging findings [2, 3]. Different terminology, level of detail, and subjective descriptions may lead to inconsistent reporting. This lack of uniformity can result in miscommunications, delays in patient management, and even diagnostic errors [4]. Additionally, the increasing workload burden on radiologists contributes to fatigue, cognitive errors, and longer report turnaround times [5, 6].

Large language models (LLMs) have emerged as a promising tool for automated radiology reporting. They can promote consistent, structured, and comprehensible reports [7, 8]. However, LLMs also introduce challenges, including biased outputs, hallucinations, and ethical and legal concerns [9, 10]. Ensuring fidelity and trustworthiness of LLM-generated reports is imperative if these models are to be safely integrated into clinical practice.

The aim of this study was to systematically review the current literature on the use of LLMs in generating radiology reports. We discuss their benefits, limitations, and potential future implications.

## Methods

### Literature Search

We systematically searched the literature to identify studies describing the generation of radiology reports using LLMs. We searched MEDLINE, Google Scholar, Scopus, and Web of Science for papers published from January 2015 up to February 2025. The full search process including Boolean operators is detailed in the **Supplementary Materials**. In addition, we checked the reference lists of selected publications and the “Similar Articles” feature in PubMed, to identify additional publications. Ethical approval was not required, as this is a systematic review of previously published research and does not include individual participant information. Our study followed the Preferred Reporting Items for Systematic Reviews and meta-analyses (PRISMA) guidelines [11]. The study is registered with PROSPERO (CRD 42025647882)

### Study Selection

All search results were imported into a single CSV table and deduplicated. Two authors (YA and VS) independently screened titles and abstracts for relevance. Potentially eligible articles were retrieved in full text and assessed by YA and VS. Discrepancies were resolved by a third author (EK).

### Inclusion and Exclusion Criteria

Included were full-text peer-reviewed publications in English focusing on radiology report generation using LLMs. We excluded non-English articles, non-original research, non-peer reviewed, studies that did not assess LLMs, and studies that did not explicitly assessed radiology reports or impressions generation. **Figure 1** presents the flow diagram of the screening and inclusion process.

**Figure 1.**
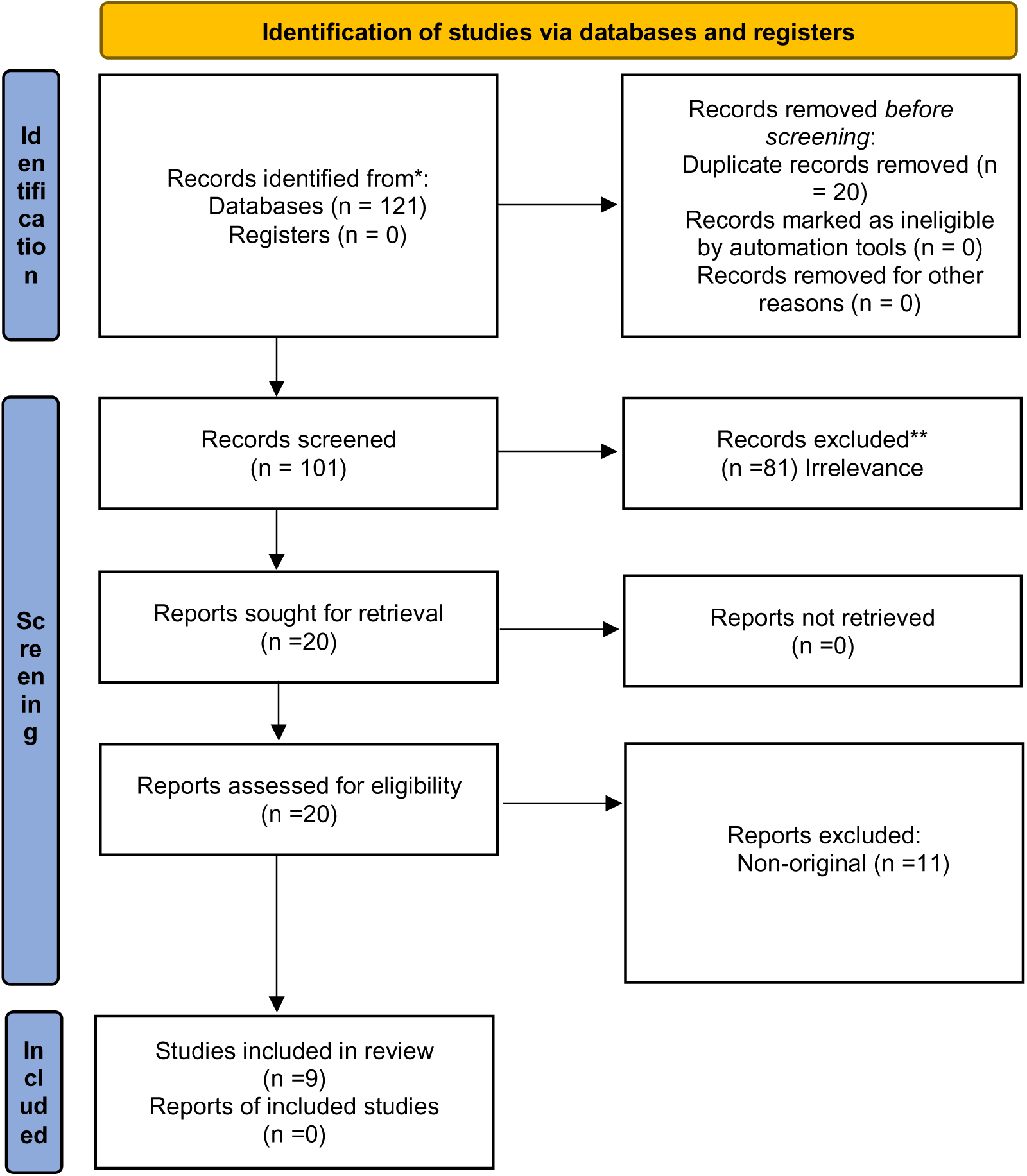
Flow diagram of inclusion and exclusion process

### Quality Assessment

The risk of bias and applicability was evaluated using the tailored Quality Assessment of Diagnostic Accuracy Studies tool 2 (QUADAS-2). (**Figure 2**.)

**Figure 2.**
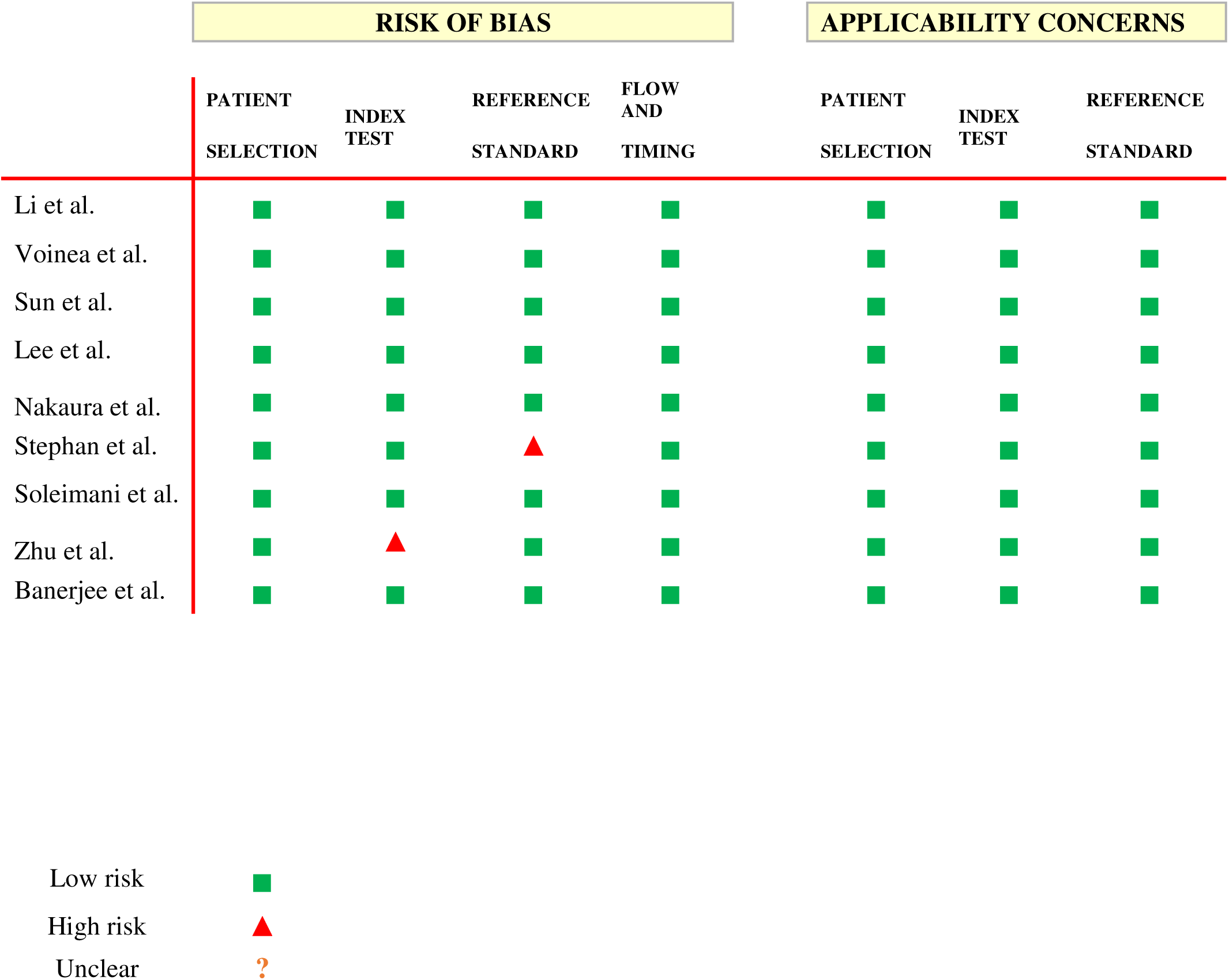
Risk of bias and applicability assessment using QUADAS-2 tool

### Performance Evaluation

LLM performance involved either quantitative or qualitative approaches. Quantitative evaluations assessed similarity between AI-generated and reference reports. These were based on automatic natural language processing (NLP) metrics [12, 13, 14]. Explanations and key differences of these metrics are detailed in **Supplementary Table 1.** Qualitative metrics were based on human evaluation.

## Results

### Study Selection and Characteristics

Nine studies were included in this review, published between September 2023 and August 2024. Six studies utilized different versions of generative pre-trained transformers (GPT) (66%). Five studies focused on chest X-ray (CXR) reports (55%). One study focused on breast ultrasound reports (11%), one used MRI reports (11%), one used dental X-ray images (11%), and two used CT reports (22%) (**Table 1**). The results of the studies are summarized in **Table 2**. **Figure 3** provides an overview of the characteristics of the included studies.

**Figure 3.**
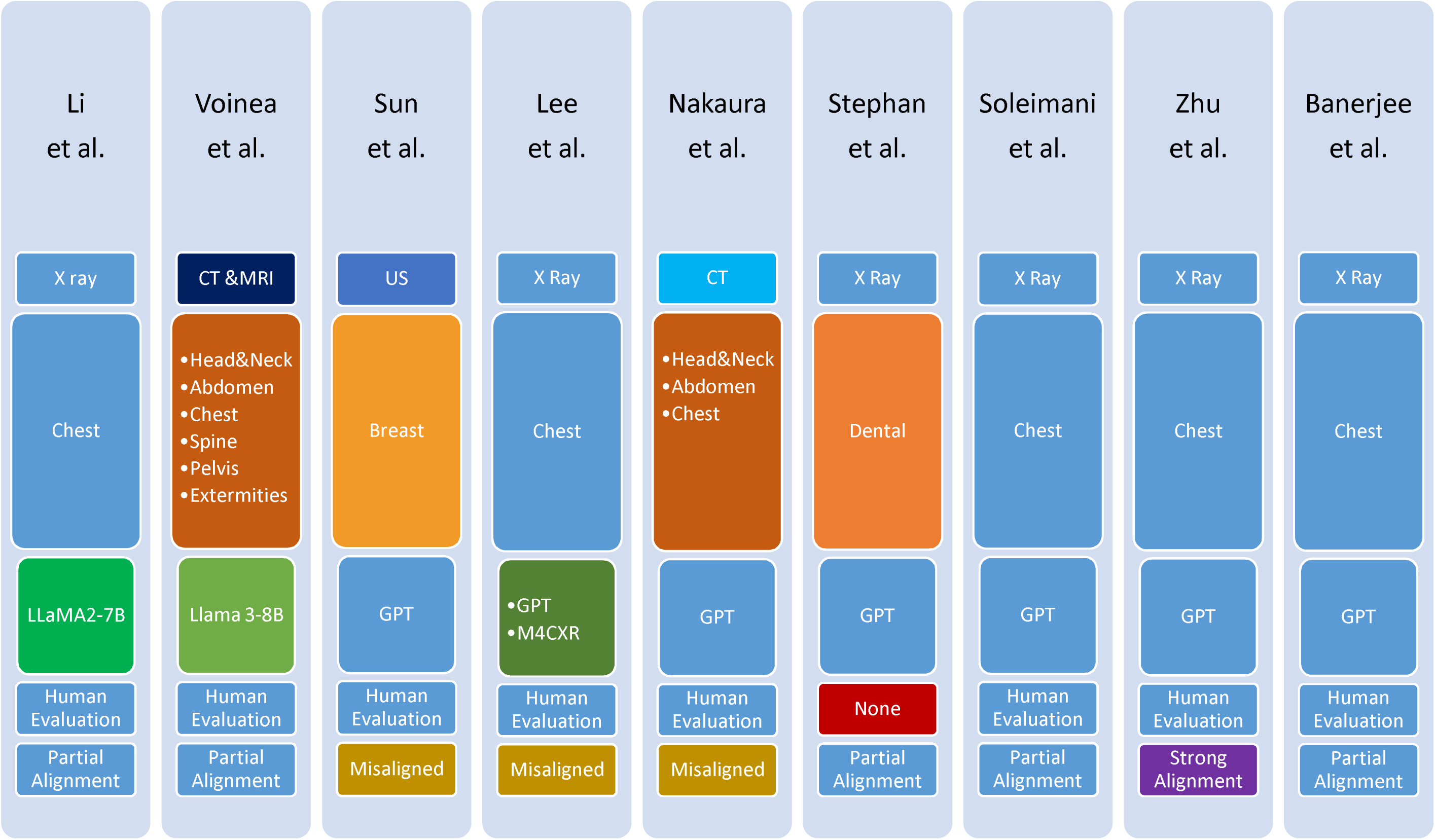
Overview of differences and similarities across studies

**Table 1.** General features of reviewed studies.

| Study | Dataset | LLM | Modality | Alignment |
| --- | --- | --- | --- | --- |
| **Li et al.** <sup>[15]</sup> | IU-Xray<br>MIMIC-CXR | LLaMA2-7B | Chest X-Ray | Partially aligned<br>(outperformed<br>existing models) |
| **Voinea et al.** <sup>[16]</sup> | 21,152 Radiology<br>Reports from the<br>University of<br>Medicine and<br>Pharmacy of<br>Craiova's Imaging<br>Center | Llama 3-8B | MRI<br>CT | Partially aligned<br>(acceptable<br>structure, minor<br>inconsistencies) |
| **Sun et al.** <sup>[17]</sup> | 131 Ultrasound<br>Reports from Beijing<br>Friendship Hospital | GPT-3.5 | Breast<br>Ultrasound | Misaligned<br>(misclassification<br>errors) |
| **Lee et al.** <sup>[18]</sup> | 826 PA Chest<br>Radiographs from<br>Inha University<br>Hospital, Incheon | M4CXR<br>GPT-4o | Chest X-Ray | Misaligned due<br>to hallucinations |
| **Nakaura et al.** <sup>[19]</sup> | 28 Images from the<br>Radiology Review<br>Manual, 8th ed | GPT-2<br>GPT-3.5<br>GPT-4 | CT | Misaligned<br>(missed<br>diagnostic<br>details) |
| **Stephan et al.** <sup>[20]</sup> | Unknown<br>Orthopantomogram<br>(OPG) | GPT<br>(Version not<br>mentioned) | Dental X-Ray | Well-aligned in<br>readability, but<br>lacked<br>diagnostic depth |
| **Soleimani et al.**<br><sup>[21]</sup> | 1000 Records from<br>MIMIC-CXR | GPT<br>(Version not<br>mentioned) | Chest X-Ray | Partially aligned<br>in impression<br>generation |
| **Zhu et al.** <sup>[22]</sup> | MIMI C-CXR | GPT-4<br>GPT-3.5 | Chest X-Ray | Well-aligned<br>with expert<br>assessments |
| **Banerjee et al.**<br><sup>[23]</sup> | Private Dataset<br>Containing Chest X-ray Reports<br>Worldwide | GPT-4 | Chest X-Ray | FineRadScore-GPT-4 was well-aligned, but<br>other metrics<br>failed |

**Table 2.** Evaluation metrics and key results.

| Study | Task | Human Evaluation | BLEU | BERTScore | ROUGE | METEOR | Clinical Metrics | Other |
| --- | --- | --- | --- | --- | --- | --- | --- | --- |
| Li et al. <sup>[15]</sup> | Full Report Generation | Expert radiologists (No. Not mentioned) | BLEU-4: **0.140** | High (Exact score not mentioned) | ROUGE-L improved | Best Scores in MIMIC-CXR (**0.165**) and IU-Xray (**0.218**) | RadGraph-F1, RadCliQ, CheXpert Similarity (Best scores) | CIDEr used |
| Voinea et al. <sup>[16]</sup> | Full Report Generation | 13 Doctors | score of **0.225** | Precision: **0.8054**<br>Recall: **0.7710**<br>F1: **0.786** | ROUGE-1: **0.4998**<br>ROUGE-L: **0.4628** | **0.4282** (+0.0561) | N/A | **Turing-like Evaluation:**<br>21.8% of cases preferred AI<br>**Overall rating:**<br>3.65 out of 5 |
| Sun et al. <sup>[17]</sup> | Impression Generation | 57 Doctors | N/A | N/A | N/A | N/A | BI-RADS Classification ($\kappa = 0.68$)<br>Malignancy Diagnosis ($p = 0.033$) | **Turing Test:**<br>35.34% of AI reports mistaken for human-written |
| Lee et al. <sup>[18]</sup> | Chest X-Ray Interpretation | Two board-certified radiologists | N/A | N/A | N/A | N/A | M4CXR Accuracy: 60.5%<br>GPT-4 Accuracy: 42.5% | Higher hallucination rates in GPT-4 |
| Nakaura et al. <sup>[19]</sup> | Differential Diagnosis | One board-certified radiologist<br>One radiology resident | N/A | N/A | N/A | N/A | Top-1 Accuracy: GPT-4 **54%**, GPT-3.5 **54%**, GPT-2 **21%**<br>Radiologists: **100%** | Kappa analysis: Overall quality ($\kappa = 0.42$) |
| Stephan et al. <sup>[20]</sup> | Dental Radiology Reporting | Dental Experts<br>Radiology instructors | N/A | Precision: **0.967**<br>Recall: **0.958**<br>F1: **0.962** | N/A | N/A | N/A | **Readability** (FRE: $p = .49$<br>LIX: $p = .09$)<br>**Findings Per Spreadsheet**<br>AI included fewer findings ($P = .003$ and $P = .014$). |
| Soleimani et al. <sup>[21]</sup> | Findings & Impression Generation | N/A | N/A | **Findings Similarity:**<br>Bart **99.3%**, | N/A | N/A | N/A | Cosine Similarity, TF-IDF, Jaccard, Sequence |
|  |  |  |  | XLM **98.9% Impression Similarity:** Word-Embedding **84.4%**, MPNet **75.7%** |  |  |  | Matching, XLM, MPNet |
| Zhu et al. <sup>[22]</sup> | Full Report Evaluation | Six radiologists | Lower correlations | N/A | Lower correlations | METEOR ($\tau = 0.29$) | CheXbert, RadCliQ, RadGraph-F1, FineRadScore-GPT-4 | Few shot GPT-4 (5-shot) performed best ($\tau = 0.48$) |
| Banerjee et al. <sup>[23]</sup> | Metric Evaluation | Internal medicine attending physician<br>Radiology resident | BLEU-2 (worst performance, negative correlation) | BERTScore highly sensitive to stylistic differences | N/A | N/A | FineRadScore-GPT-4 (best metric, no stylistic bias) | RadGraph-F1, CheXpert, RadCliQ |

Non-peer-reviewed publication were excluded from our analysis, but their results are detailed in the **Supplementary Material,** along with A summary presented in **Supplementary Table 2.**

### LLM Performance in Generating Full Radiology Reports (Image-to-Text) vs. Radiology Impressions

Six studies evaluated full report generation based on image analysis. Three studies focused on radiology impression generation based on the findings sections of the reports (**Figure 4**).

**Figure 4.**
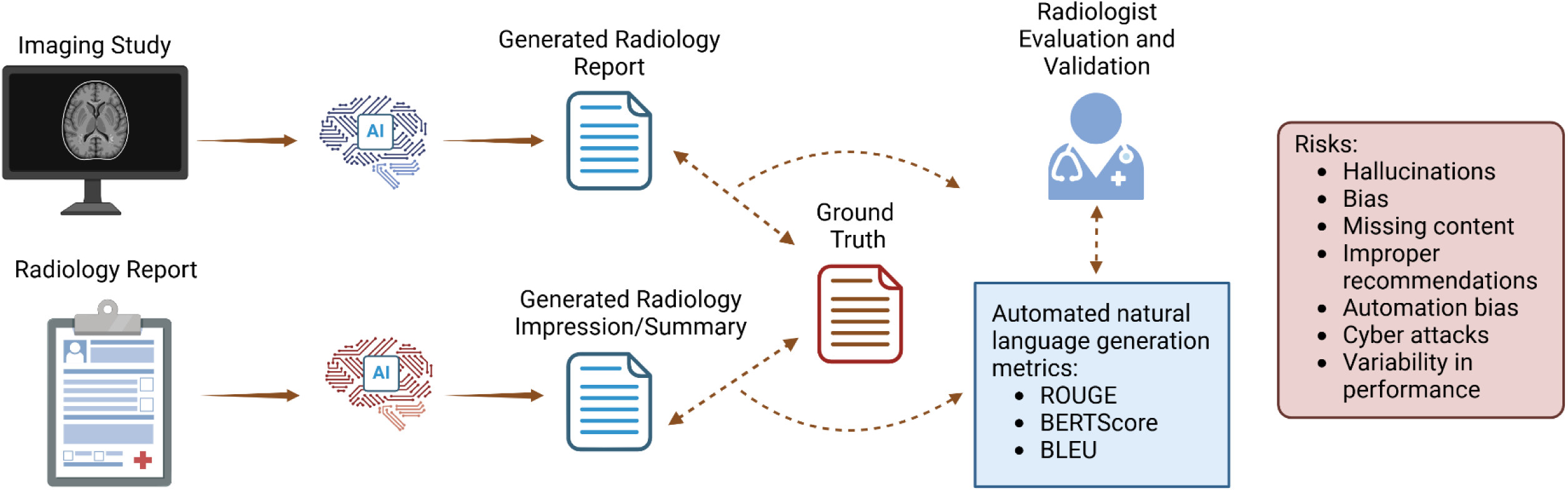
Schematic illustration demonstrating AI report generation

Li et al. [15] showed that an LLM enhanced with a knowledge graph outperformed base models in generating radiology reports. They extracted regional image features using a transformer model. These were then combined with disease-specific features from the knowledge graph. Their model outperformed other pre-trained LLMs for radiology reporting. However, its reliance on a constrained knowledge graph limited generalization beyond the scope of chest X-rays and introduced potential biases.

Voinea et al. [16] assessed the fine-tuned Llama 3-8B for full reports generation. Their model produced clinically relevant impressions achieving a ROUGE-1 F1 score of 0.4998, ROUGE-L F1 score of 0.4628, and a BERTScore F1 of 0.4282. While AI-generated conclusions were preferred in 21.8% of cases, radiologists still found that in many other cases, the model’s conclusions lacked clinical judgment and exhibited biases due to dataset limitations. Similarly, Sun et al. [17] evaluated GPT-3.5’s ability to generate BI-RADS classifications and treatment recommendations from breast ultrasound reports, noting structured output but frequent misdiagnoses, overdiagnoses, and classification inconsistencies (**Table 2**, **Table 3**).

**Table 3.** Limitations and drawbacks of AI performance.

| Study | Limitations |  |  |  |  |
| --- | --- | --- | --- | --- | --- |
| Li et al. <sup>[15]</sup> | Limited Scope of the Knowledge Graph | Focus on Chest X-rays Only | Multimodal Fusion Complexity | Dependency on Knowledge Graph Quality | Potential Bias from Training Data |
| Voinea et al. <sup>[16]</sup> | Dataset Limitations | Dependence on Input Quality | Lack of Full Clinical Context | Challenges in Complex Cases | Voinea et al. <sup>[14]</sup> |
| Sun et al. <sup>[17]</sup> | Lower Accuracy in BI-RADS Classification and Malignancy Diagnosis | Inconsistencies Between Report Components | Overdiagnosis of Benign Lesions | Text-Only Input Limitation | Reduced Performance on Complex Cases |
| Lee et al. <sup>[18]</sup> | Lower Diagnostic Accuracy for GPT4o | Higher Rate of False Findings for GPT4o | Inferior Performance in Location Accuracy for GPT4o | Higher Hallucination Rate for GPT4o | Slower Processing Time for GPT4o |
| Nakaura et al. <sup>[19]</sup> | Lower accuracy in differential diagnosis | Incomplete alignment with radiologists' impressions | Hallucination and fabrication of findings | Moderate interobserver agreement |  |
| Stephan et al. <sup>[20]</sup> | Omission of Critical Diagnostic Information | Challenges in Interpreting Complex Odontogram Data | Dependence on Prompt Design | Potential for Bias Due to Training Data | Variability in Readability |
| Soleimani et al. <sup>[21]</sup> | Limited Understanding of Context and Subtle Findings | Inconsistent Performance Across Report Sections | Lack of Ground Truth Interpretation Process | Potential Overfitting to Report Style | Lack of Image Analysis Capability |
| Zhu et al. <sup>[22]</sup> | Lower Performance of GPT-3.5 Compared to GPT-4 | Stylistic Sensitivity and Bias in Traditional Metrics | Limited Clinical Insight in NLG and CE Metrics | Challenges in Capturing Sentence-Level Nuances | Dependence on Annotated Data from Experts |
| Banerjee et al. <sup>[23]</sup> | Sensitivity to Stylistic Differences | Poor Alignment with Expert Judgment | Inconsistent Performance Across Sites |  |  |

Lee et al. [18] compared M4CXR and GPT-4 for interpreting chest X-rays and generating associated radiology reports. M4CXR outperformed GPT-4 in diagnostic accuracy and anatomical localization, while GPT-4 produced more false findings and hallucinations. Nakaura et al. [19] showed that GPT-3.5 and GPT-4 achieved high readability in their generated reports, but underperformed in differential diagnoses compared to radiologists.

Stephan et al. [20] found that GPT-generated reports from dental panoramic radiographs (OPGs) had high textual similarity and better readability than student-written reports, but lacked critical diagnostic details.

### Fine-Tuned vs. Out-of-the-Box Model Performance

There were six studies evaluating base LLMs, and three studies evaluated pretrained or fine-tuned models. Li et al. [15] showed that a fine-tuned LLM outperformed base models on NLP metrics (BLEU-4 improved by 4.5%, higher RadGraph-F1, BERTScore, and RadCliQ) (**Table 2**). Additionally, the fine-tuned model demonstrated improved alignment with human reports as measured by NLP metrics, though direct human evaluation was not reported. Soleimani et al. [21] reported that GPT models generated structurally coherent reports but missed subtle findings that radiologists identified. Voinea et al. [16] showed that a fine-tuned Llama 3-8B significantly improved BLEU, ROUGE, METEOR, and BERTScore over base models for full radiology report generation.

Fine-tuning generally improved performance. For example, Zhu et al. [22] showed that a fine-tuned GPT-4 achieved higher correlation with expert evaluations (τ = 0.48) compared to base models. Regressed GPT-4, referring to a post-processing approach where a regression model is trained using expert-annotated radiology reports to adjust GPT-4-generated scores, improved alignment further (τ = 0.64). However, fine-tuned models still exhibited hallucinations and inconsistent clinical accuracy when lacking diverse training data (**Table 2**, **Table 3**).

### Automated Metrics vs. Human Expert Evaluation

Banerjee et al. [23] examined text-based evaluation metrics, finding that most automated metrics overemphasized stylistic similarity rather than clinical correctness. The study highlighted FineRadScore-GPT-4 as the only metric that correlated well with expert assessments, primarily due to its focus on clinical relevance and semantic accuracy rather than mere context similarity (**Table 2**).

Banerjee et al. [23] also noted that Natural Language Generation (NLG) metrics, which evaluate textual similarity and fluency using measures such as BLEU, ROUGE, and METEOR, correlated poorly with expert assessments, often ranking stylistically similar AI reports higher, regardless of diagnostic accuracy. Specifically, they assessed BLEU-2.

Zhu et al. [20] proposed a detailed few shots (5) GPT-4 evaluation model, which outperformed traditional metrics in aligning with expert radiologists. Metrics like FineRadScore-GPT-4 were better aligned with radiologists’ evaluations, while metrics like RadGraph F1 and RadCliQ [24] helped capture clinical relevance (**Table 2**).

### Key Limitations of Current AI in Radiology Reports

Across the nine studies reviewed, AI-generated reports exhibited hallucinations (reported in 44% of studies, 4/9), misdiagnoses (55%, 5/9), and missing clinical details (66%, 6/9) as frequent errors. Lee et al. [18] reported that GPT-4 misidentified anatomical locations and over-reported findings. Stephan et al. [20] highlighted AI’s struggles with integrating broader clinical context. Sun et al. [17] found GPT-3.5 frequently overdiagnosed benign lesions, and Nakaura et al. [20] showed that GPT models underperformed in differential diagnosis accuracy (**Table 3**). Examples of the various prompts used in the studies are detailed in **Table 4**. Limitations examples mentioned in the studies are presented in **Figure 5 and Figure 6**. Further examples are included in the **Supplementary Material.**

**Figure 5.**
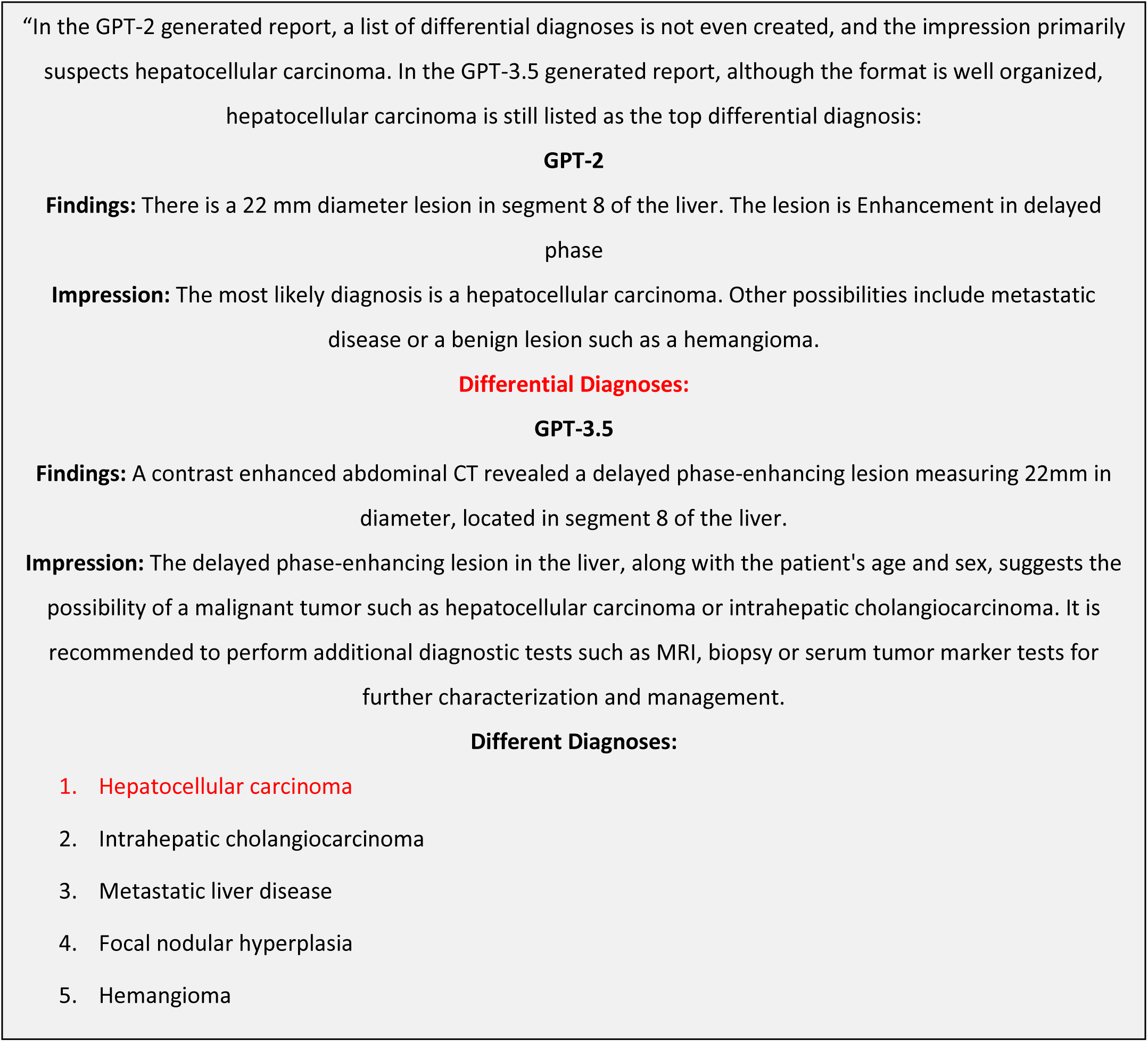
Limitation Example, lower accuracy in differential diagnosis, Nakaura et al.

**Figure 6.**
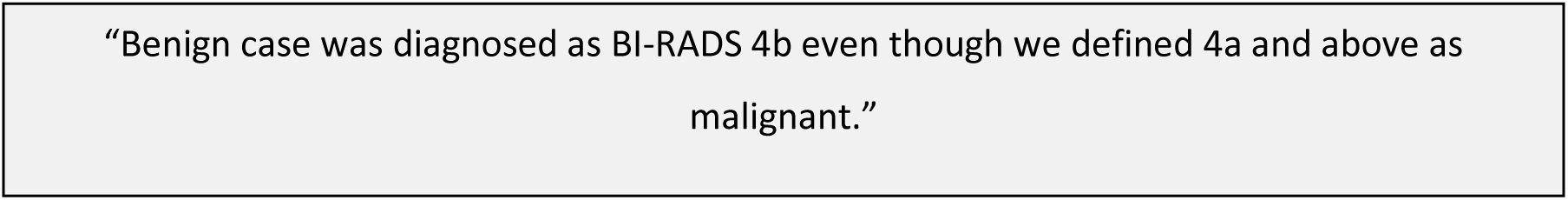
Limitation Example, lower accuracy in BI-RADS classification and malignancy diagnosis, Sun et al.

**Table 4.** Various prompt examples.

| Study | Prompt Examples |
| --- | --- |
| Li et al. <sup>[15]</sup> | "Generate a comprehensive and detailed diagnosis report for this radiology image." |
| Voinea et al. <sup>[16]</sup> | N/A |
| Sun et al. <sup>[17]</sup> | "Please summarize the BI-RADS classification, benign/malignant, and brief treatment recommendations based on the following ultrasound diagnostic results. The following is a description of breast ultrasound:<br>"Examination findings:<br>Both breasts have uneven echogenicity and disordered structure, with no mass or abnormal blood flow signals observed in the left breast.<br>In the right breast, multiple low-to-no echogenic nodules are found, with the largest one located at the 3 o'clock direction about 0.9cm from the nipple in the glandular layer, measuring approximately 0.7x0.3cm, having clear and regular boundaries, and no evident blood flow signals within.<br>No abnormally enlarged lymph nodes are detected in both axillae." |
| Lee et al. <sup>[18]</sup> | "This is a chest PA image. Tell me more about what is going on?" |
| Nakaura et al. <sup>[19]</sup> | "...Provide a concise and prioritized catalogue of differential diagnoses without a corresponding disease description, comprising no more than five suspects, ranked by level of suspicion." |
| Stephan et al. <sup>[20]</sup> | "Please write a dental radiology report for the following findings derived from an orthopantomogram using appropriate radiological terminology: [Findings list provided here]. Structure the report into a findings section and an impression section. Please follow the style and conventions of radiological reports used in dental education." |
| Soleimani et al. <sup>[21]</sup> | "Utilizing the prognoses outlined in Step 1 and the findings acquired in Step 2, compose a radiology report comprising two sections that accurately describe the health condition of the patient who visited the hospital. Please respond only with the information I request. Please provide the information as follows: Section One: Offer an impression in a single paragraph. Section Two: Suggest a diagnosis concisely, without any additional recommendations." |
| Zhu et al. <sup>[22]</sup> | "Your task is to evaluate the prediction sentence by sentence, comparing it with the original report. You should also have one overall score from 0-5 for prediction sentences with respect to the original report based on your subjective impression. And I only need the csv format for the result." |
| Banerjee et al. <sup>[23]</sup> | "Pretend you are a radiologist and format the content of these notes in a polished findings and impressions section. Your findings section may be long or short. Your impression should only have 1-3 lines. If you are unsure about an abbreviation, term, or other odd phrasing, make your best guess. Match |
|  | the style of this radiology report..." |

## Discussion

In this systematic review, we explored the role of LLMs in radiology report generation. Several studies showed that these models can improve consistency, readability, and structure. However, they also reveal critical limitations in diagnostic accuracy, clinical reasoning, and attention to nuanced clinical context, where human radiologists demonstrate clear superiority.

Producing a meaningful radiology report requires technical accuracy, clear communication, and clinical synthesis [25]. It must be understandable and actionable for various stakeholders. These include patients, clinicians, and support staff. Notably, none of the included studies examined how clinicians or patients perceived the AI-generated reports, although LLMs could potentially offer clearer, more accessible communication [26, 27]. A recurring theme across the reviewed studies was the variability in LLMs’ performance across different tasks. Stylistic bias and overfitting to training data patterns compromised both performance and the validity of the evaluation metrics. Several studies demonstrated that automated metrics, such as BLEU, ROUGE, and METEOR, were disproportionally influenced by stylistic similarities, rather than clinical accuracy [15, 16, 23]. Scores that rely on semantic similarities, such as BERTScore and RadGraph may offer more relevant insights, but these also fall short of alignment with actual clinical relevancy [28]. Human evaluations, on the other hand, are subjective and not scalable for large datasets. This creates a gap in reliable metrics for evaluation.

Several studies also noted the importance of the input quality [16, 17, 20]. This dependence was particularly evident in studies involving multimodal data, such as breast ultrasound or chest X-ray interpretation, where imaging findings alone were insufficient for comprehensive reporting [16, 17, 21]. Collaboration between AI and radiologists may be a promising direction. In studies where radiologists actively refined or corrected AI outputs, report quality improved. Yet, such approaches are only practical if they reduce workload rather than increase the burden of validation and correction in the clinical routine [29].

Future research should focus on improving evaluation frameworks, including diverse datasets to mitigate biases, and prospectively validating AI-generated reports in clinical workflows. Real-world validation remains sparse but essential to translate these advances into reliable clinical tools. Numerous commercial radiology reporting software tools are already widely used in radiology practice. Yet their performance, failure modes, and reliability are largely unstudied in the academic literature. Understanding how these systems compare to newer, research-driven models will be essential. Furthermore, with the pace of generative AI development, it remains to be determined how base models compare to LLMs that are fine-tuned on specialized radiology datasets.

Fine-tuning can align a model’s output with a specific reporting style. This can improve user satisfaction and clinical relevance [30]. However, this customization may also potentially increase the risk of “automation bias”, where radiologists trust these familiar outputs without adequate review. Fine-tuning also exposes training data to adversarial attacks [31]. It also demands resources, including large, high-quality datasets and specialized expertise. Meanwhile, proprietary models continue to improve rapidly, potentially offering more advanced clinical reasoning without explicit domain-specific training [32]. Another factor is test-time computation. That is the computational resources used when the model generates a report. Iterative refinement can potentially yield higher-quality outputs but at greater cost and longer processing times [33, 34]. As such, both fine-tuning and test-time computations will have to be weighed against clinical needs, and resource availability.

Our review has several limitations. Due to heterogeneity in study design and data, a meta-analysis was not performed. The majority of the studies focused on one modality (X-ray) and included limited LLM performance in other imaging modalities, such as CT or MRI. One study did not include a human expert evaluation of the LLMs’ performance. We excluded non-English studies, which can limit diverse insight into LLM performance in a global context. Also, most of the studies did not include examples of the AI limitations, which could provide insight into real-world integration issues. None of the studies included have evaluated these tools in a real-world hospital workflow. Additional studies in a real-world environment will be needed to explore AI fidelity in radiology reporting.

In conclusion, LLMs may be a valuable tool for radiology reporting, but their responsible deployment requires careful attention to clinical accuracy, transparency, and human oversight. Ensuring alignment between AI and human goals is essential. By integrating AI as an adjunct, future systems can benefit from the efficiency and consistency of LLMs while preserving the clinical reasoning and contextual understanding unique to human expertise.

## Data Availability

All data produced in the present work are contained in the manuscript

## Declarations

### Ethics approval and consent to participate

Not applicable

### Consent for publication

Not applicable

### Availability of data and materials

All data generated or analyzed during this study are included in this published article

### Authors’ contributions

YA conducted the literature search, data synthesis and wrote the primary manuscript. BSG, JC, and PK reviewed and edited the manuscript. GNN, EK, and VS contributed to the conceptualization, data synthesis, and editing of the manuscript. All authors read and approved the final manuscript.

## Acknowledgements

Not applicable

