## Supplementary material for "Large Language Models in Radiology Reporting—A Systematic Review of Performance, Limitations, and Clinical Implications": Supplementary Material.docx

### Literature Search Process

The search was conducted using the following Boolean operators:

("artificial intelligence" OR "AI" OR "machine learning" OR "deep learning" OR "natural language processing" OR "NLP" OR "large language models" OR "LLM")

AND

("alignment" OR "AI alignment" OR "value alignment" OR "goal alignment" OR "model alignment" OR "ethical alignment" OR "robustness" OR "interpretability" OR "explainability")

AND

("radiology" OR "radiological" OR "radiological reports" OR "imaging reports" OR "diagnostic imaging" OR "medical imaging" OR "computed tomography" OR "CT" OR "magnetic resonance imaging" OR "MRI" OR "ultrasound" OR "X-ray")

AND

("reporting" OR "clinical reporting" OR "automated reporting" OR "AI-generated reports" OR "structured reporting" OR "text generation" OR "documentation" OR "narrative reports")

### Supplementary Table 1**.** Metric Types and differences

| Metric | Type | Purpose | Key Difference |
| --- | --- | --- | --- |
| BLEU | NLG | Measures n-gram precision between AI and reference text | More precision-focused; emphasizes word-for-word matching |
| BLEU-2 | NLG | Measures bigrams overlap between AI and reference text | Focuses on exact matching of 2-word sequences; lacks semantic understanding |
| ROUGE | NLG | Measures overlapping sequences between AI and reference text | Primarily recall-focused, assessing how much of the reference text is captured |
| ROUGE-L | NLG | Focuses on longest common subsequence overlap | Captures structural similarity, useful for longer texts |
| BERTScore | NLG | Measures semantic similarity using contextual embeddings | Captures meaning beyond exact word overlap by using pre-trained embeddings |
| METEOR | NLG | Combines exact matches, synonyms, and stemming for similarity | Balances precision, recall, and meaning, more flexibility |
| CIDEr | NLG | Weighs term frequency to emphasize rare but important terms | Prioritizes rare medical terms to highlight unique findings |
| RadGraph-F1 | Clinical | Assesses clinical entity and relation extraction accuracy | Evaluates how well AI extracts structured medical information from reports |
| RadCliQ-v0/1 | Clinical | Combines clinical correctness and quality into a composite score | Assesses both medical content accuracy and overall text coherence |
| CheXpert | Clinical | Evaluates agreement between AI and radiologist for 14 predefined chest findings | Uses CheXpert labeler to classify findings; focused on chest X-rays |
| CheXbert Similarity | Clinical | Evaluates similarity in structured finding labels between AI and human reports | Focused more on similarity rather than exact label matching. |
| FineRadScore-GPT-4 | Hybrid (NLG + Clinical) | Evaluates textual similarity and clinical correctness, using GPT-4 to rate reports | Uses GPT-4 to combine NLG fluency with domain-specific clinical relevance. |

### Limitations Examples

#### Banerjee et al.

***Inconsistent Performance Across Sites***

“Reports from Germany were automatically translated to English, resulting in atypical wording choices (e.g. “delineable”, “ateal”).”

#### Sun et al.

***Inconsistencies Between Report Components***

“a report indicated a benign diagnosis but suggested a biopsy”

#### Nakaura et al.

***Hallucination and fabrication of findings***

“GPT-4.0 report is overall quite similar to a human-generated report, and the differential diagnosis is reasonable. However, it includes information that was not input, such as calcification and cystic degeneration:

A contrast enhanced CT scan of the brain shows a 49 mm enhancing mass in the vicinity of the sella turcica. The mass extends both above and below the diaphragma sellae, occupying the suprasellar and intrasellar regions. The mass compresses the optic chiasm and displaces the pituitary gland inferiorly. There is no evidence of calcification, cystic changes, or bony reaction.

A contrast enhanced CT scan of the brain shows a 49 mm enhancing mass in the vicinity of the sella turcica. The mass extends both above and below the diaphragma sellae, occupying the suprasellar and intrasellar regions. The mass compresses the optic chiasm and displaces the pituitary gland inferiorly. There is no evidence of calcification, cystic changes, or bony reaction.”

***Lower accuracy in differential diagnosis***

“In the GPT-2 generated report, the possibility of a renal tumor accompanied by surrounding edema is low, and a list of differential diagnoses is not even created. In the GPT-3.5 generated report, although the lesion is located in the kidney, the differential diagnoses include adrenal adenoma”

**GPT 2**:

**Finding**: There is an approximately 35 mm mass with low-density fatty content present in the inferior portion of the right renal lobe. The mass is well margined and has no obvious necrosis or calcification. There is minimal surrounding edema.

**Impression**: Low-density fatty mass in the right kidney, suggestive of a benign lesion such as a complex cystic renal mass or angiomyolipoma. Differential diagnosis also includes liposarcoma or lipoma, which may be considered given the well-circumscribed nature of the mass. Additional workup with MRI or PET/CT may be necessary to further characterize the mass and guide management.

**Differential Diagnosis**:

**GPT 3:**

**Findings:** A contrast-enhanced CT study was performed on a 75-year-old male patient, revealing the presence of a 35mm fat-containing renal mass located in the inferior portion of the right renal organ.

**Impression:** The observed fat-containing renal mass could be indicative of angiomyolipoma, a benign tumor typically composed of adipose tissue, smooth muscle, and blood vessels. However, given the patient’s age and the size of the mass, a malignant tumor such as renal cell carcinoma cannot be entirely ruled out. Further evaluation with biopsy or MRI may be recommended to confirm the diagnosis.

**Differential Diagnosis:** 1. Angiomyolipoma 2. Renal cell carcinoma 3. Liposarcoma 4. Adrenal adenoma 5. Wilms’ tumor

***Hallucination and fabrication of findings***

“In the GPT-4.0 generated report, the overall quality is quite good; however, there is a description of “calcification” in the image findings, which was not part of the input information:

**GPT 4**:

**Finding**: Contrast-enhanced CT of the kidneys shows a fat-containing mass in the inferior portion of the right kidney, measuring 35 mm in diameter. The mass has a heterogeneous appearance with areas of soft tissue attenuation and calcification

**Impression**: The imaging findings are most consistent with a renal angiomyolipoma, which is a benign tumor that contains blood vessels, smooth muscle cells, and fat cells. Renal angiomyolipomas are the most common fat-containing renal mass and are usually asymptomatic unless they cause hemorrhage or compression of adjacent structures. Other less likely differential diagnoses include renal cell carcinoma, which is a malignant tumor that may contain macroscopic fat and calcification, and renal lipoma or liposarcoma, which are rare tumors that contain mostly fat

**Differential Diagnosis**: Renal angiomyolipoma (high suspicion) - Renal cell carcinoma (low suspicion) - Renal lipoma or liposarcoma (very low suspicion)

### Non-Peer-Reviewed Publications

Pellegrini et al. [1] developed RaDialog, a vision-language model designed for radiology report generation and interactive diagnostic assistance. The system integrates radiologist-provided observations in a structured format with an LLM, enabling both automated reporting and real-time collaboration between radiologists and AI. The model was trained on a semi-automatically labeled dataset for chest X-ray interpretation, and using parameter-efficient fine-tuning.

The model’s performance was evaluated using a combination of automated natural language processing (NLP) metrics and human expert assessments. The model achieved a BERTScore of 9.5, outperforming comparison models in semantic similarity. It also demonstrated higher text coherence with a BLEU-4 score of 0.4, a METEOR score of 14.0, and a ROUGE-L score of 27.1, all of which were superior to baseline models. Human expert evaluation further validated the model's performance, with radiologists rating RaDialog-generated reports as more clinically accurate

Liu et al. [2] introduced HC-LLM, a historical-constrained LLM framework aimed at improving radiology report generation by incorporating historical imaging data to capture disease progression. They extracted both time-shared and time-specific features from longitudinal chest X-rays. HC-LLM achieved higher BLEU-4 (0.142), ROUGE-L (0.287), and METEOR (0.162) scores, outperforming baseline models and maintaining high diagnostic accuracy even in cases where historical data was absent during testing.

Zeng et al. [3] focused on impression generation. They proposed a multi-agent LLM system. Their framework employed three specialized agents: a retrieval agent that identifies relevant prior reports, a radiologist agent responsible for generating impressions, and a reviewer agent that refines outputs based on quality feedback.

The model demonstrated improvements in NLG metrics, achieving BLEU-4 of 24.22, ROUGE-L of 0.4364, and BERTScore of 0.7434, outperforming single-agent approaches. Additionally, qualitative evaluation performed by GPT-4 indicated an 8.5% increase in coherence and a 6.2% improvement in clinical relevance scores compared to single-agent baselines.

Chen et al. [4] introduced a large vision-language model tailored for radiology reporting based on 3D CT scans, particularly chest CTs. The model was evaluated using public and private datasets, with results demonstrating superior performance in diagnostic accuracy and textual coherence compared to both vision transformer-based and CNN-based models.

3D-CT-GPT achieved a BLEU score of 0.2323, ROUGE-1 of 0.3008, ROUGE-2 of 0.0706, and ROUGE-L of 0.1567, outperforming M3D, which scored 0.0869, 0.1336, 0.0227, and 0.1028 respectively. Additionally, an ablation study demonstrated that fine-tuning the CT-ViT model improved performance, with BLEU increasing from 0.2950 to 0.3476, ROUGE-1 from 0.4163 to 0.4446, METEOR from 0.3037 to 0.3198, and BERTScore_F1 from 0.8809 to 0.8862.

### Supplementary Table 2. Summary of Non-Peer Reviewed Publications

| Study | Aim | Method | Key Results |
| --- | --- | --- | --- |
| Pellegrini et al. | Developed RaDialog, a vision-language model for radiology report generation and interactive diagnostic assistance. | Evaluated performance using NLP metrics (BLEU, ROUGE, METEOR, BERTScore) and radiologist assessments. | RaDialog achieved BLEU-4 score of 0.4, a METEOR score of 14.0, and a ROUGE-L score of 27.1, all of which were superior to baseline models. Radiologist assessments confirmed better interactive diagnostic support. |
| Liu et al. | Introduced HC-LLM, a historical-constrained LLM integrating longitudinal imaging data for improved radiology report generation. | Incorporated historical imaging data to extract time-shared and time-specific features for chest X-rays; evaluated using NLP and diagnostic metrics. | HC-LLM achieved higher BLEU-4 (0.142), ROUGE-L (0.287), and METEOR (0.162) scores demonstrating accuracy even without historical data. |
| Zeng et al. | Proposed a multi-agent LLM framework for impression generation, incorporating retrieval, radiologist, and reviewer agents. | Compared single-agent vs. multi-agent models using NLP metrics and GPT-4 qualitative assessment. | Multi-agent LLM outperformed single-agent models, achieving BLEU-4 of 24.22, ROUGE-L of 0.4364, and BERTScore of 0.7434, outperforming single-agent approaches |
| Chen et al. | Developed 3D-CT-GPT, a vision-language model designed for automatic radiology report generation from 3D chest CT scans. | Benchmarked against existing models (M3D) on public and private datasets; assessed using BLEU, ROUGE, METEOR, and BERTScore. | 3D-CT-GPT surpassed M3D with BLEU 0.2323 vs. 0.0869, ROUGE-1 0.3008 vs. 0.1336, and METEOR 0.3198 vs. 0.3037. |
